# Forecasting the spread of COVID-19 in Nigeria using Box-Jenkins Modeling Procedure

**DOI:** 10.1101/2020.05.05.20091686

**Authors:** Rauf Rauf Ibrahim, Hannah Oluwakemi Oladipo

**Affiliations:** Bioresources Development Center, National Biotechnology Development Agency, Ilorin Nigeria; Department of mathematics, University of Abuja, Nigeria

## Abstract

**Objective:** This study is focused on the analysis of the spread of Covid-19 in Nigeria, applying statistical models and available data from the NCDC. We present an insight into the spread of Covid-19 in Nigeria in order to establish a suitable prediction model, which can be applied as a decision-supportive tool for assigning health interventions and mitigating the spread of the Covid-19 infection.

**Methodology:** Daily spread data from February 27 to April 26, 2020, were collected to construct the autoregressive integrated moving average (ARIMA) model using the R software. Stability analysis and stationarity test, parameter test, and model diagnostic were also carried out. Finally, the fitting, selection and prediction accuracy of the ARIMA model was evaluated using the AICc model selection criteria.

**Results:** The ARIMA (1,1,0) model was finally selected among ARIMA models based upon the parameter test and Box–Ljung test. A ten-day forecast was also made from the model, which shows a steep upward trend of the spread of the COVID-19 in Nigeria within the selected time frame.

**Conclusion:** Federal Government of Nigeria through the presidential task force can apply the forecasted trend of much more spread to make more informed decisions on the additional measures in place to curb the spread of the virus. Application of the model can also assist in studying the effectiveness of the lockdown on the on the spread of Covid-19 in Nigeria.

## Introduction

COVID-19 has been spreading rapidly globally, with a considerable impact on global morbidity, mortality and healthcare utilization. This viral disease is caused by the Coronaviruses (CoV) which belong to the genus ‘coronavirus’ of the Coronaviridae family. All CoVs are pleomorphic RNA viruses characterized by crown-shape (the name “coronavirus” is derived from the Greek Κορώνα, meaning crown.) peplomers with 80-160 nM in size and the genome of CoV contains a linear, single-stranded RNA molecule of positive (mRNA) polarity and about 28-32Kb in length (Sahin, 2020; Woo, Huang, Lau, & Yuen, 2010). Coronaviruses possess the largest genomes among all identified RNA viruses, and the large genome gives the virus extra plasticity in accommodating and modifying genes. Hence high recombination rates are observed in CoVs due to constantly developing transcription errors, and RNA dependent RNA polymerase (RdRP) jumps (Woo et al., 2010). Coronaviruses are zoonotic pathogens, found in humans and various animals with the ability to exhibit clinical features in human. In humans, there are four prototypic human CoV (hCoV) that cause endemic and epidemic respiratory disease, including the human alpha coronaviruses 229E and NL63 and the human beta coronaviruses OC43 and HKU1 (Drexler et al., 2010). Infected patients can be asymptomatic or symptomatic affecting the gastrointestinal, respiratory, hepatic and neurological systems and may result in hospitalization at the intensive care unit (Drexler et al., 2010; Woo et al., 2010; Yin & Wunderink, 2018).

The history of the pathogenicity of CoV is dated back to 2002 and 2003 when they were considered to be highly pathogenic due to severe acute respiratory syndrome (SARS) in Guangdong state of China for the first time. Before this outbreaks, the two most common CoV known to cause mild infections in people with the adequate immune system includes CoV OC43 and CoV 229E (Peiris et al., 2003; Yin & Wunderink, 2018). About ten years after SARS, the highly pathogenic CoV, Middle East Respiratory Syndrome Coronavirus (MERS-CoV) emerged in the Middle East countries (Zaki, Van Boheemen, Bestebroer, Osterhaus, & Fouchier, 2012) and recently in December 2019, a novel Coronavirus (nCoV) emerged at a livestock/Seafood Market in the Wuhan State of Hubei Province in China and has evolved into a global pandemic (“Seven days in medicine,” 2020). Chinese authorities announced the isolation of this novel virus on January 7 2020, the virus was named COVID-19 by the world health organization on January 12, 2020, and as at February 12, 2020, a total of 43,103 confirmed cases and 1,018 deaths had been reported (Organization, 2020).

During the ongoing pandemic, some research publications have focused on the epidemiology, trend analysis and forecasting for different cities and countries. These studies presented long-term and short-term trend using time series data from relevant database and offered forecasting applications using models such as ARIMA model, Exponential Smoothing methods, SEIR model and Regression Model.

Applying purely data-driven statistical method, Yang et al., (2020) estimated the case fatality rate (CFR) for COVID-19 in three clusters: Wuhan city, other cities of Hubei province, and other provinces of mainland China. A simple linear regression model was applied to estimate the CFR from each cluster. The result obtained showed that CFR during the first weeks of the epidemic ranges from 0.15% (95% CI: 0.12-0.18%) in mainland China excluding Hubei through 1.41% (95% CI: 1.38-1.45%) in Hubei province excluding the city of Wuhan to 5.25% (95% CI: 4.98-5.51%) in Wuhan. Their results conclusively indicate CFR of COVID-19 is lower than the previous coronavirus epidemics caused by SARS-CoV and Middle East respiratory syndrome coronavirus (MERS-CoV). To study the epidemic trend of COVID-19 in mainland China, Hubei province, Wuhan city and other provinces outside Hubei from January 16 to February 14, 2020, Zhu et al., (2020) generated the epidemic curve of the new confirmed cases, multiple of the new confirmed cases for period-over-period, multiple of the new confirmed cases for fixed-base, and the period-over-period growth rate of the new confirmed cases using data from National Health Commission. From January 16 to February 14, 2020, the cumulative number of new confirmed cases of COVID-19 in mainland China was 50 031, including 37 930 in Hubei province, 22 883 in Wuhan city and 12,101 in other provinces outside Hubei. The peak of the number of new confirmed cases in other provinces outside Hubei was from January 31 to February 4, 2020, and the peak of new confirmed cases in Wuhan city and Hubei province was from February 5 to February 9, 2020. The number of new confirmed cases in other provinces outside Hubei showed a significant decline (23% compared with the peak) from February 5 to February 9, 2020, while the number of new confirmed cases in Wuhan city (30% compared with the peak) and Hubei Province (37% compared with the peak) decreased significantly from February 10 to February 14, 2020. Fanelli & Piazza, (2020) analyzed the temporal dynamics of COVID-19 outbreak in China, Italy and France with the timeframe of January 22 to March 15 2020. A first analysis of simple day-lag maps points to some universality in the epidemic spreading and the analysis of the same data within a simple susceptible-infected-recovered-deaths model indicates that the kinetic parameter that describes the rate of recovery appears to be the same, regardless of the country, while the infection and death rates appear to be more variable. The model places the peak in Italy around March 21, 2020, with a peak number of infected individuals of about 26000 (not including recovered and dead) and a number of deaths at the end of the epidemics of about 18,000. Since the confirmed cases are believed to be between 10 and 20% of the real number of individuals who eventually get infected, the apparent mortality rate of COVID-19 falls between 4% and 8% in Italy, while it appears substantially lower, between 1% and 3% in China. Piccolomini & Zama, (2020) also proposed the modification of the Susceptible-Infected-Exposed-Recovered-Dead (SEIRD) differential model for the analysis and forecast of the COVID-19 spread in some regions of Italy. They introduced a time-dependent transmitting rate and reported the maximum infection spread for the three Italian regions firstly affected by the COVID-19 outbreak (Lombardia, Veneto and Emilia Romagna). Danon, Brooks-Pollock, Bailey, & Keeling, (2020) applied an existing national-scale metapopulation model to capture the spread of CoVID-19 in England and Wales. They captured data from population sizes and population movement, together with parameter estimates from the current outbreak in China and were able to predict the peak of the outbreak after person-person transmission was established in England and Wales. Jit et al., (2020) applied exponential growth model to fit critical care admissions from multiple surveillance to study likely COVID-19 case numbers and progress in the United Kingdom from February 16 – March 23, 2020. They estimated that on 23 March, there were 102,000 (median; 95% credible interval 54,000 - 155,000) new cases and 320 (211 - 412) new critical care reports, with 464,000 (266,000 – 628,000) cumulative cases since February 16.

On February 27, 2020, Nigeria recorded its first case of Covid-19. The index case was an Italian citizen who arrived Nigeria via the Murtala Muhammed International Airport, Lagos at 10pm aboard Turkish airline from Milan, Italy.

On March 9, 2020, the second case of Covid-19 was reported in Nigeria, this was a contact of the index case, and as the days and weeks progressed, the number of confirmed cases of Covid-19 increased stemming from both local transmission and importation from other countries. As of March, 29 2020, the total confirmed cases within Nigeria had risen to 97 (ninety-seven) and had recorded its first death on March 23 2020. The federal government on March 29, 2020, announced a lockdown on Lagos, Ogun states and the Federal Capital Territory (FCT) with effect from 11 pm of March 30, 2020. Lagos State is the epicentre of the disease and Ogun state being its boarder state while FCT had the second highest of confirmed Covid-19 cases in the country. The government had also imposed travel restriction into the country for travellers from China, Italy, Iran, South Korea, Spain, Japan, France, Germany, the US, Norway, the UK, Switzerland and the Netherlands on the March 8, 2020. It expanded these restrictions on March 21, 2020 as the nation closes its two main international airports in Lagos and Abuja. The country also suspended rail services on March 23, 2020.

As the number of cases grew nationwide in Nigeria and local transmission surged relative to the number of imported cases, there is a need to focus on more local measures to decrease the spread of COVID-19. In addition to the lockdown, social distancing rule has been enforced by cancelling mass gatherings, closing businesses except for providers of essential goods and services such as food and pharmaceutical entities and restriction of local travels. Using mathematical and statistical models as described in other studies above, study of the trend of the Covid-19 pandemic in Nigeria can provide critical information for responding to outbreaks and understanding the impact of strategies employed by the government in containing the spread of the disease. This present study focuses on the analysis of the trend of spread of COVID-19 in Nigeria, identification and application of befitting trend prediction models using available data from the NCDC. Overall, we were able to formulate time series models on the spread of COVID-19 in Nigeria, conduct a diagnostic check on the models formulated to determine the most suitable model, Estimate the parameters of the various models and forecast the COVID-19 spread variable.

### Methodology for Analysis and Forecasting

Data comprising confirmed, recovered, and death cases as variables; were retrieved from the Nigeria Centre for Diseases Control (NCDC) official COVID-19 site (https://covid19.ncdc.gov.ng/). The data refer to daily cases and cover the period from February 27, 2020, when the country recorded her index case until April 26, 2020. This data set included both “lab-confirmed” and “clinically diagnosed” cases. We emphasize the importance of the recovered cases, which is not covered in media as widely as the confirmed cases or the deaths. While, only the confirmed cases variable shows an exponential increase, most especially in the month of March 2020. However, the curve of recovered cases is flattened over the past 14 days of the study period.

Applying the obtained data, we adopted a non-stationary time series forecasting approach. Since the series is obtained over a relatively short period of time and also of a very high frequency (daily data), the Box-Jenken procedure (Musa, 2015; Yue, Shengnan, & Yuan, 2015) was employed. Forecasts were made using the Autoregressive Integrated Moving Average (ARIMA) model’s family. This family has shown good forecast accuracy over several forecasting competitions and is especially suitable for short series. This model encompasses the AR, MA or ARMA models in which differences have been taken and collectively called autoregressive integrated moving average models, or ARIMA models. A time-series {Y_t_} is said to follow an integrated autoregressive moving average model if the d^th^ difference *W_t_* = ∇*^d^Y_t_* is a stationary ARMA process. If {Wt} follows an ARMA (p, q) model, we say that {Y_t_} is an ARIMA (p, d, q) process. Considering the generalized form of ARIMA model, taking d = 1 or at most 2 (Kane, Price, Scotch, & Rabinowitz, 2014).

Thus, an ARIMA (p, 1, q) process. With W_t_ = Y_t_−Y_t−1_, we have

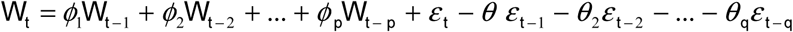

Or, in terms of the observed series,

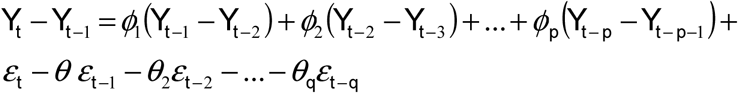

Stationary Test and Jarque-Bera Test for normality were applied as a prerequisite for the application of the ARIMA time-series model (Sato, 2013; Wu, Guan, Guo, & Zhou, 2008). Statistical approaches for model selection (such as information criteria, which measure the maximum likelihood of a model) was used to select the model that better reflect the nature of the data, thus model with the best model’s selection criterion. We produce ten-days-ahead point forecasts and prediction intervals and update our forecasts every ten days. Please note that this is not an ex-post analysis, but a real, live forecasting exercise. The R. Console 3.5.2 version statistical software was used for this study’s data analysis.

## Results

The overall distribution data of COVID-19 from February 27, 2020 to April 26, 2020, were collected and analyzed. A total of 1120 COVID-19 cases were observed from February 27, 2020 to April 26, 2020, and the incidence showed a wave-like increasing tendency day by day, with the trend of rectilinear rise after March 27, 2020, when the first lockdown was enacted by the Federal Government of Nigeria. The recovery rate as of April 26, 2020, was 16.09%, which stood for the peak incidence and recovery ratio since the index case of COVID-19 in Nigeria.

### ARIMA Model Forecasting Analysis

Sequence Characteristic Analysis and Transformation: Firstly, a daily sequence from February 27, 2020 to April 26, 2020, was calculated and its chart was drawn, as shown in Figure 3A the original sequence showed a mixed trend of an upward or downward trend with a seasonal cycle rhythm, which was not smooth and had uneven variances. The original sequence was transformed into a random one through the methods of first difference transformation and once seasonal difference successively. After that, the time sequence displayed a random and stationary trend (Figure 3B and Table 2). As is shown in Figures 3 (A-B), a total of 117 cases on April 21, 2020, represented the peak daily report of COVID-19 since the index case in Nigeria.

**Figure 1.**
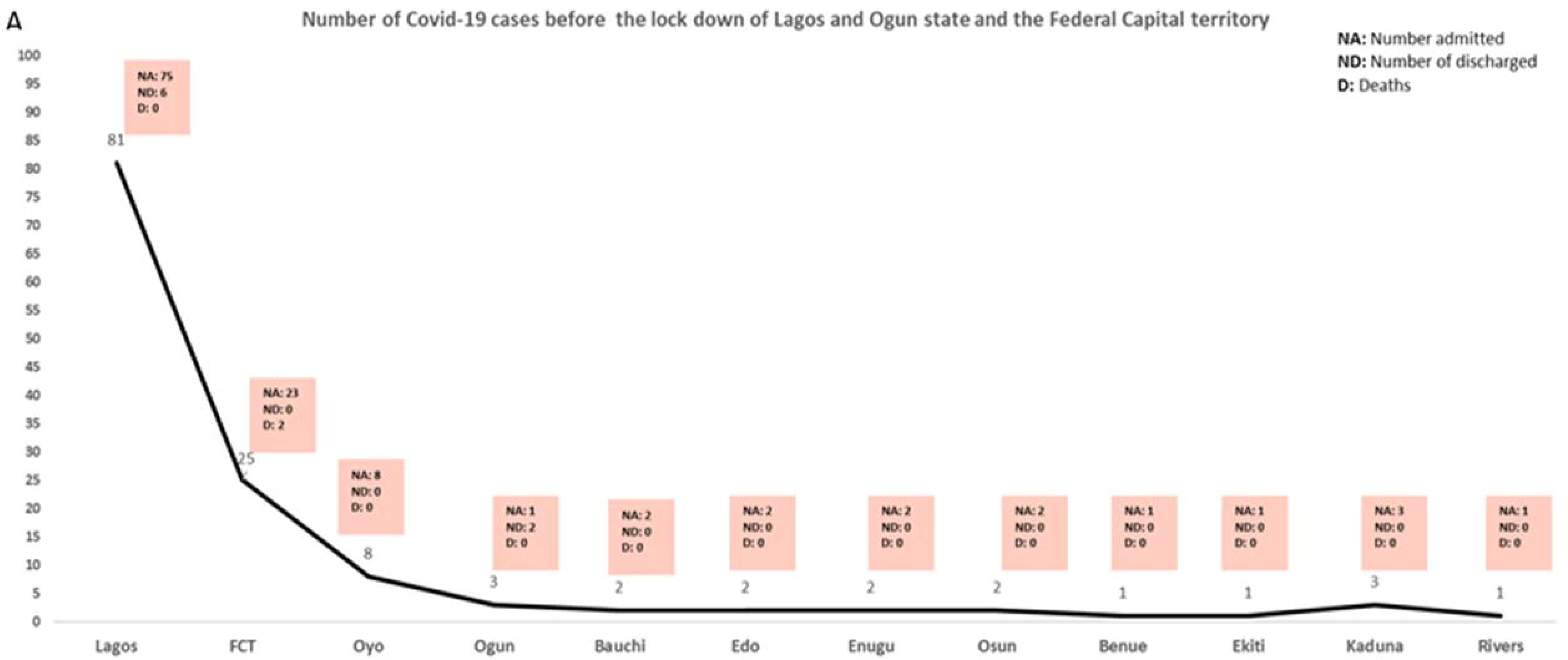
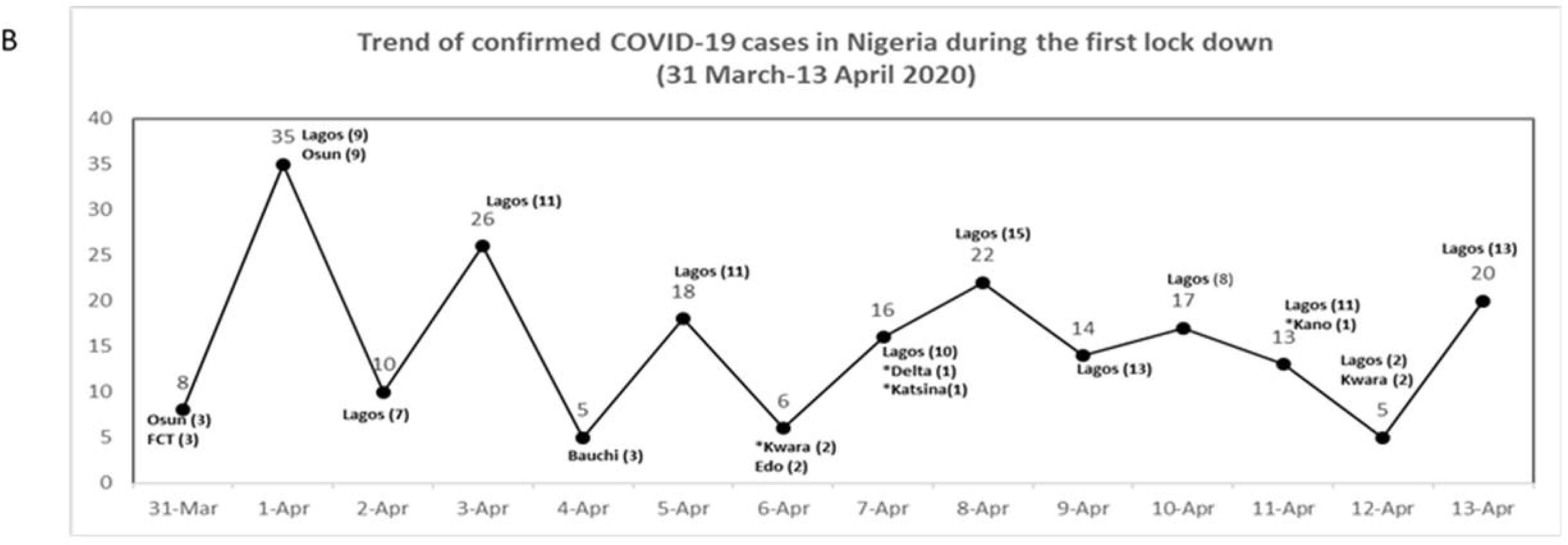
A-B. A Number of confirmed COVID-19 cases before the lock down, B Number of confirmed cases across states that already had confirmed cases and also new states (*) with confirmed cases during the innitial two weeks lockdown

**Figure 2:**
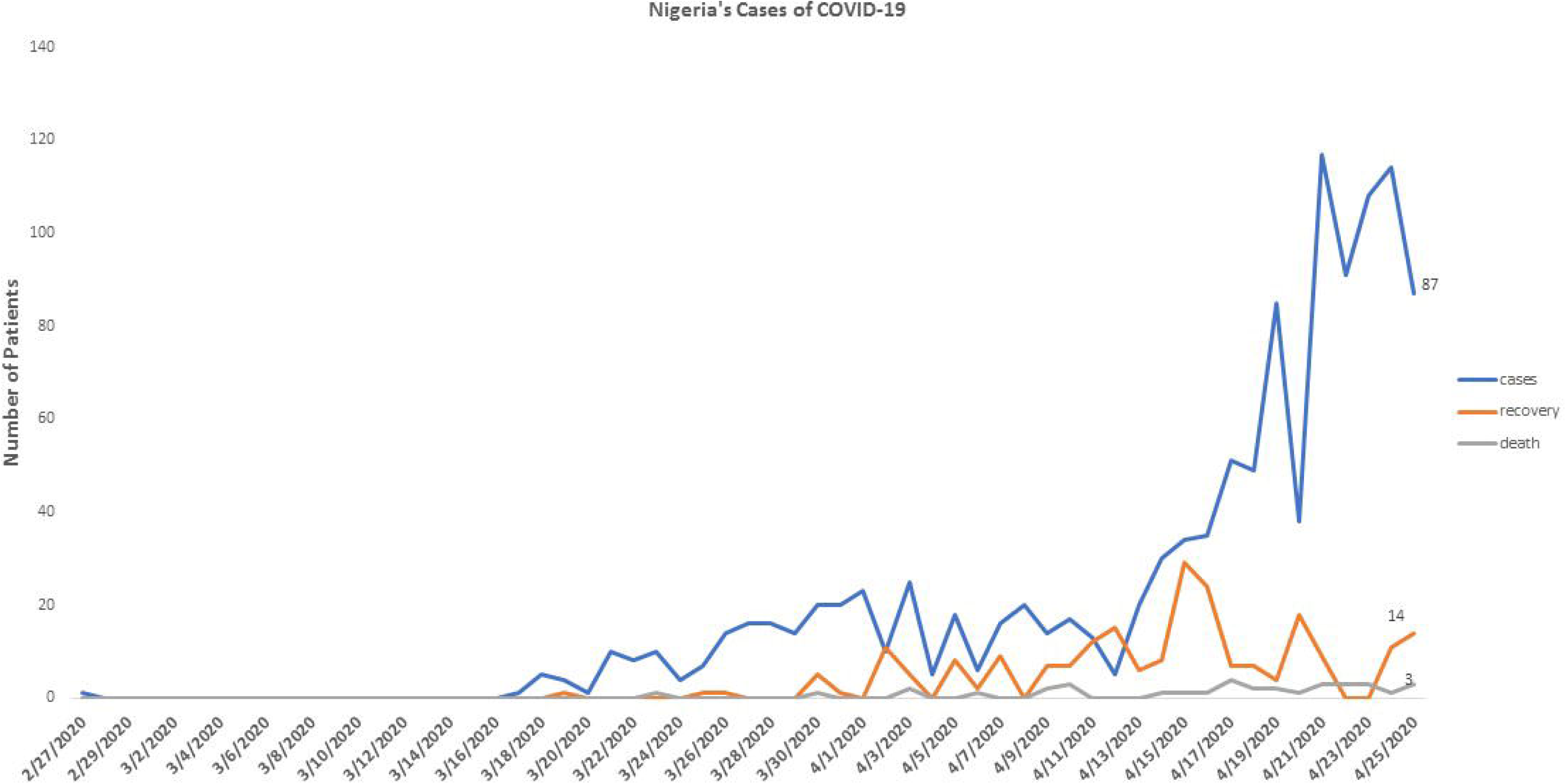
.Epidemiological Characteristics of COVID-19 in Nigeria from February 27 to April 26, 2020

**Figure 3.**
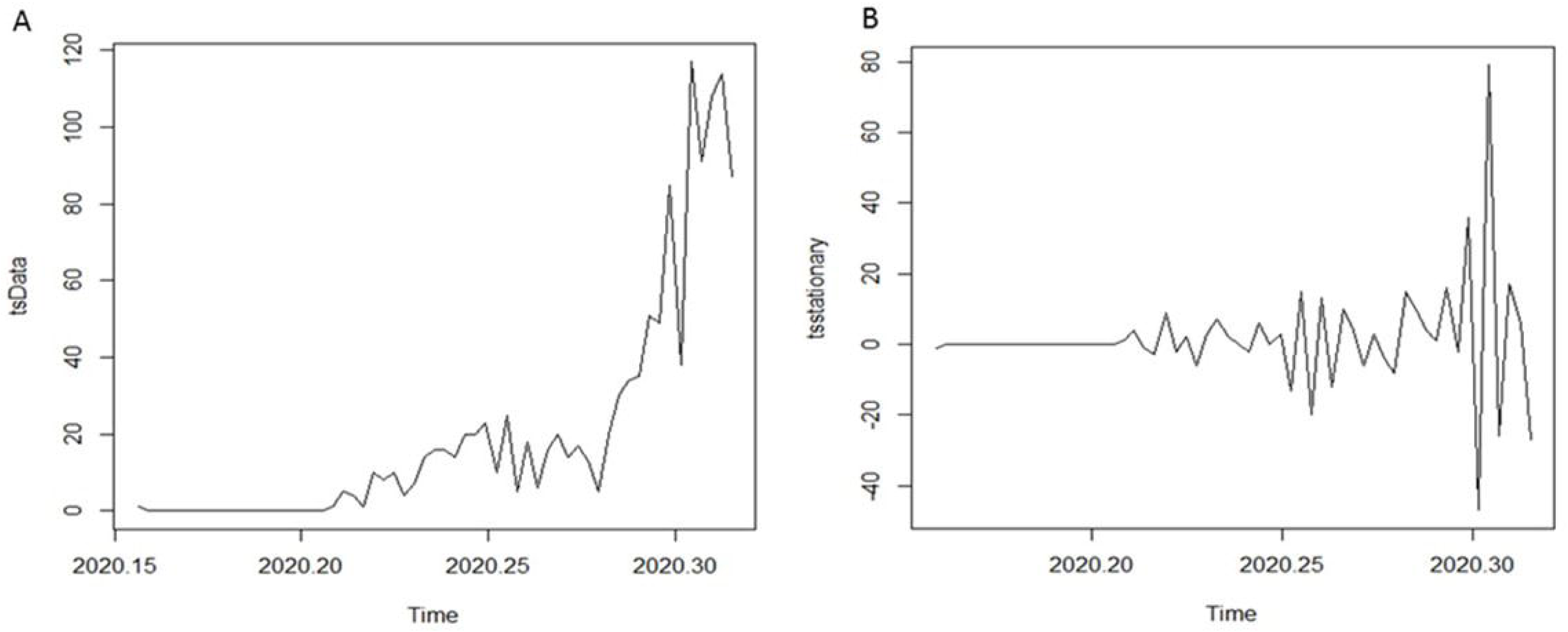
A-B (A-B) (A) Time Distribution Plot of the Daily Confirmed Cases of COVID-19 in Nigeria. (B) Time Distribution Plot of the First Differenced Series of Daily Confirmed Cases of COVID-19 in Nigeria.

**Figure 4.**
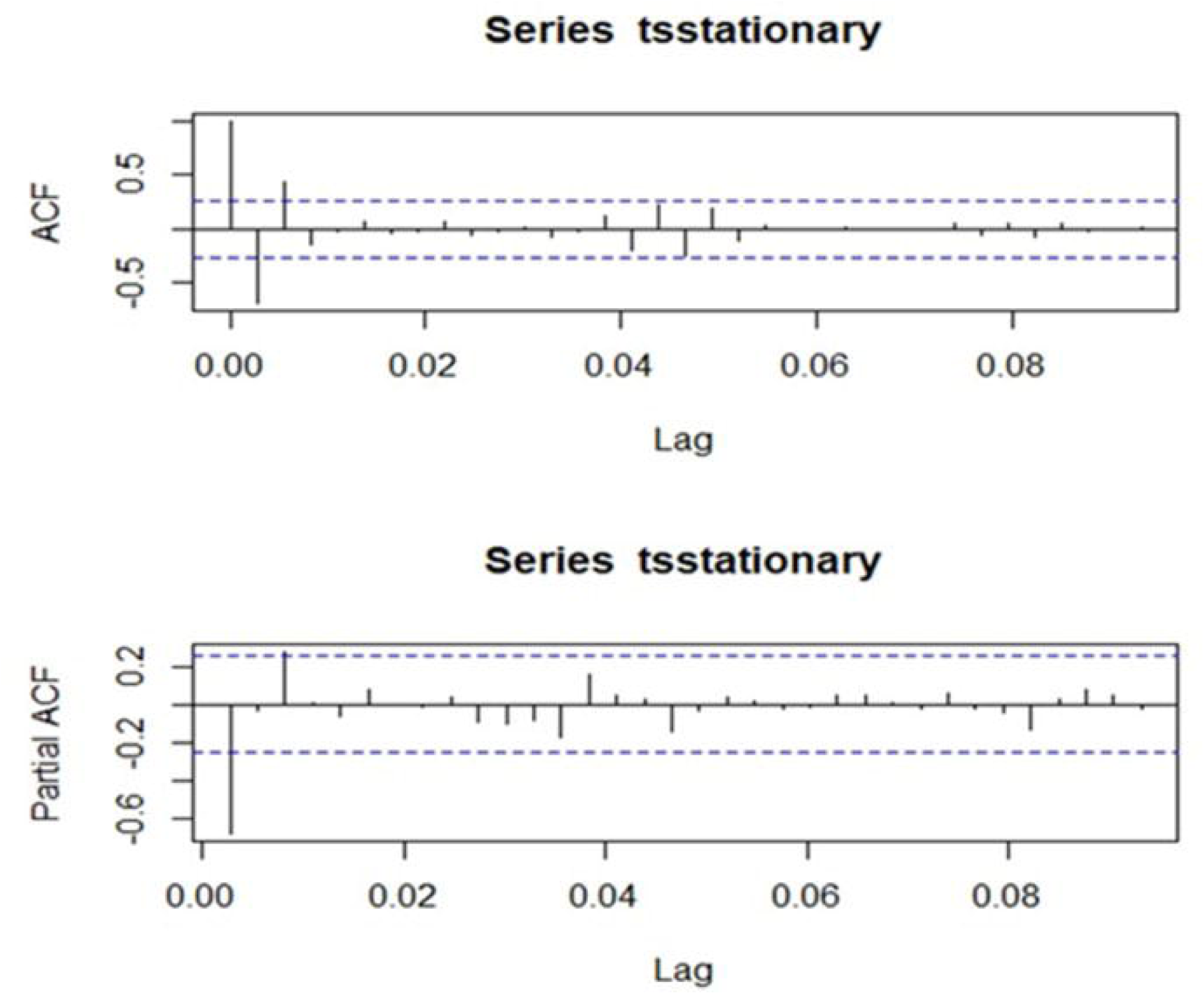
ACF and PACF plots to determine the candidate models

**Figure 5.**
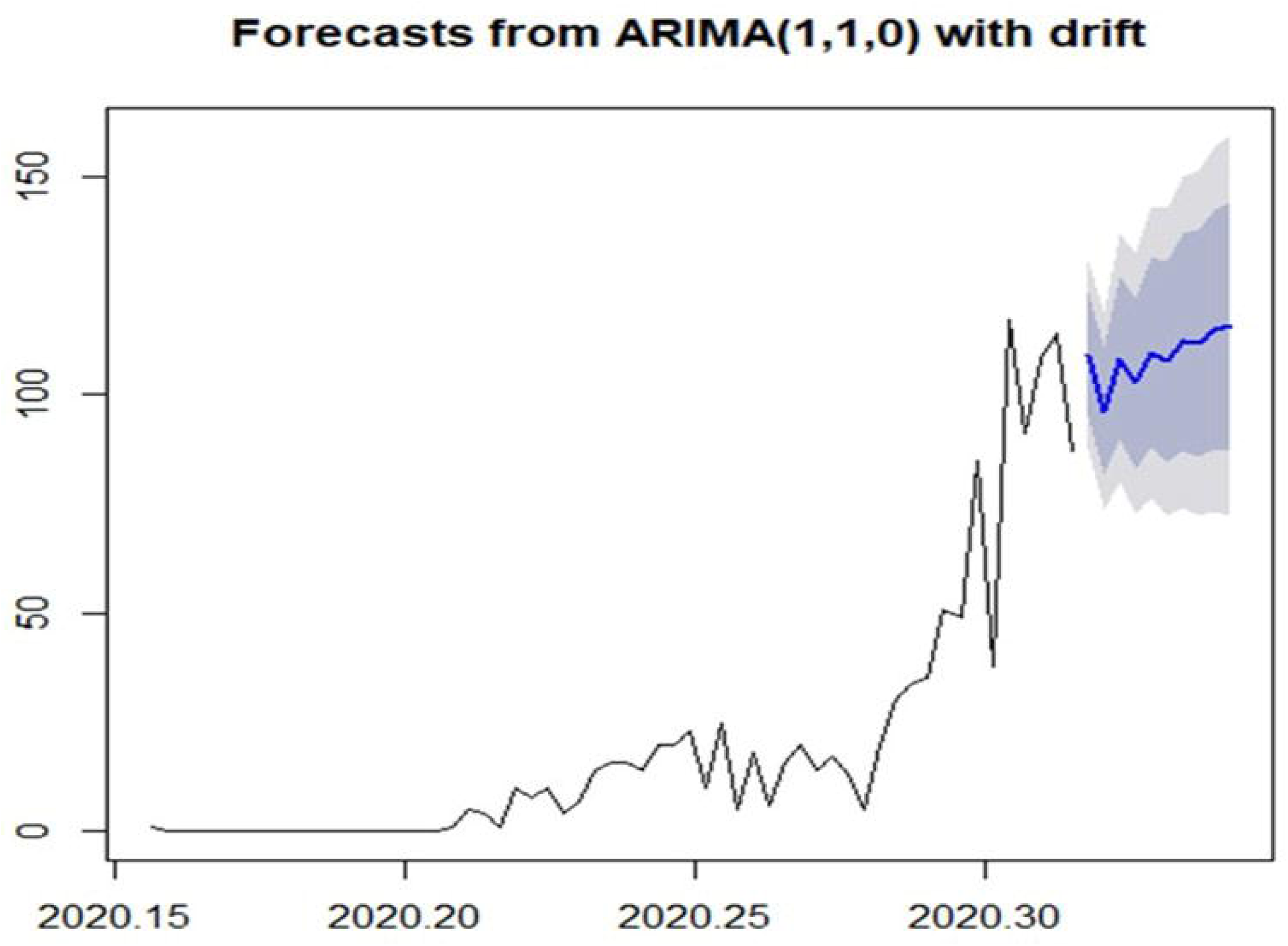
A Ten-Day Forecast of COVID-19 Spread in Nigeria

**Table 1:** Descriptive Statistics

|  | N | Range | Minimum | Maximum | Sum | Mean | Std. Deviation |
| --- | --- | --- | --- | --- | --- | --- | --- |
| Cases | 59 | 117.00 | .00 | 117.00 | 1182.00 | 20.0339 | 30.2067 |
| Recovery | 59 | 29.00 | .00 | 29.00 | 222.00 | 3.7627 | 6.3091 |
| Death | 59 | 4.00 | .00 | 4.00 | 35.00 | .5932 | 1.0524 |
| Valid N (listwise) | 59 |  |  |  |  |  |  |

**Table 2:**
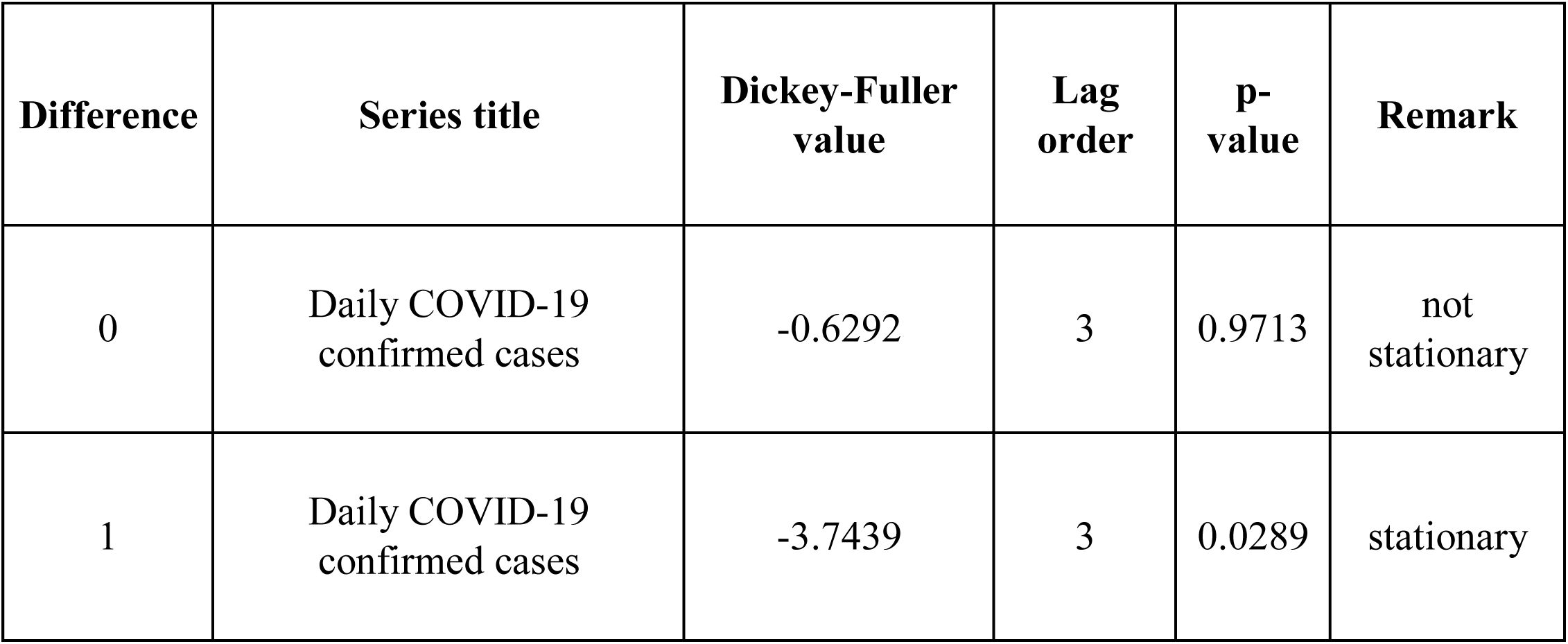
Stationarity Test of the series with Augmented Dickey-Fuller Test

| Difference | Series title | Dickey-Fuller value | Lag order | p-value | Remark |
| --- | --- | --- | --- | --- | --- |
| 0 | Daily COVID-19 confirmed cases | -0.6292 | 3 | 0.9713 | not stationary |
| 1 | Daily COVID-19 confirmed cases | -3.7439 | 3 | 0.0289 | stationary |

### Stationarity Test of the series with Augmented Dickey-Fuller Test

To effectively apply the ARIMA modelling technique, the series must be stationary and free of any form of trend. To confirm the status of the series (daily confirmed cases of covid19 in Nigeria), the ADF test was thus use to validate the stationarity observed from the series transformation (ADF test: t= −3.7439 and P <0.05) (Table 2), thus making the series obtaining stationarity at 1^st^ difference. The series was however not observed to be stationary at level, which is the natural state of the data, then we transformed the series to make stationary by first difference. This makes the series stationary and ready for modelling inline the Box-Jenken ARIMA modelling approach.

### Candidate Model Identification

A list of candidate models or potential model combinations will be obtained from the plots of the autocorrelation function (ACF) and the partial autocorrelation function (PACF). The order of the model was determined according to ACF and PACF after once common difference. Based on the spikes observed from the ACF and PACF chart, the following candidate models are formulated. The candidate model with the lowest Akaike information criterion correction (AICc) value was selected as the best model to fit the daily spread series of COVID-19 in Nigeria.

From the ACF and PACF graph (as shown in Figure 3 above) and the models trace summary table (Table 3), we were able to observe the following candidate models and also using the AICc model selection criterion, we detect that the ARIMA(1,1,0) with drift as the model with lowest AICc value.

**Table 3.** Model Fit for COVID-19 Cases in Nigeria

| SN | Candidate models | AICc | Type |
| --- | --- | --- | --- |
| 1 | ARIMA(2,1,2) | 444.9136 | with drift |
| 2 | ARIMA(0,1,0) | 479.719 | with drift |
| 3 | ARIMA(1,1,0) | 443.2092 | with drift |
| 4 | ARIMA(0,1,1) | 458.1584 | with drift |
| 5 | ARIMA(0,1,0) | 478.115 |  |
| 6 | ARIMA(2,1,0) | 445.9576 | with drift |
| 7 | ARIMA(1,1,1) | 445.2854 | with drift |
| 8 | ARIMA(2,1,1) | Inf | with drift |
| 9 | ARIMA(1,1,0) | 445.0218 |  |

### Model coefficients test

As observed from the above table, the best model of the candidate models is the ARIMA (1,1,0) base on the AICc criterion. The model is then estimated with its parameter estimates for forecasting the daily spread series of COVID19 in Nigeria.

### Model estimation and z test of coefficients

`Estimate Std. Error z value Pr(>|z|)`

`ar1    -0.713370    0.093684    -7.6146    2.645e-14     ***`

`drift    1.708284    0.838514    2.0373    0.04162    *`

`Signif. codes:    0    '***'    0.001    '**'    0.01    '*'    0.05    '.'    0.1    ' '  1`

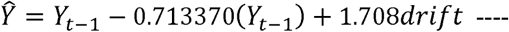 20091686 which is thus the model to forecast the spread of covid19 in Nigeria.

### Forecasting Using ARIMA Model

As shown in Figure 3, the daily spread data from April 26 to May 5, 2020, were predicted using the ARIMA(1,1,0) model based on the spread of COVID-19 in Nigeria from February 27 to April 26, 2020, the results of which suggested that the predicted values fitted well with the actual values.

The forecast Table 4 above contains the date, point forecast and the high and low confidence limits values of the forecast. The daily forecast is the point forecast but can be with the 95% confidence limit of the Hi95 and Lo95 values on the table.

**Table 4:**
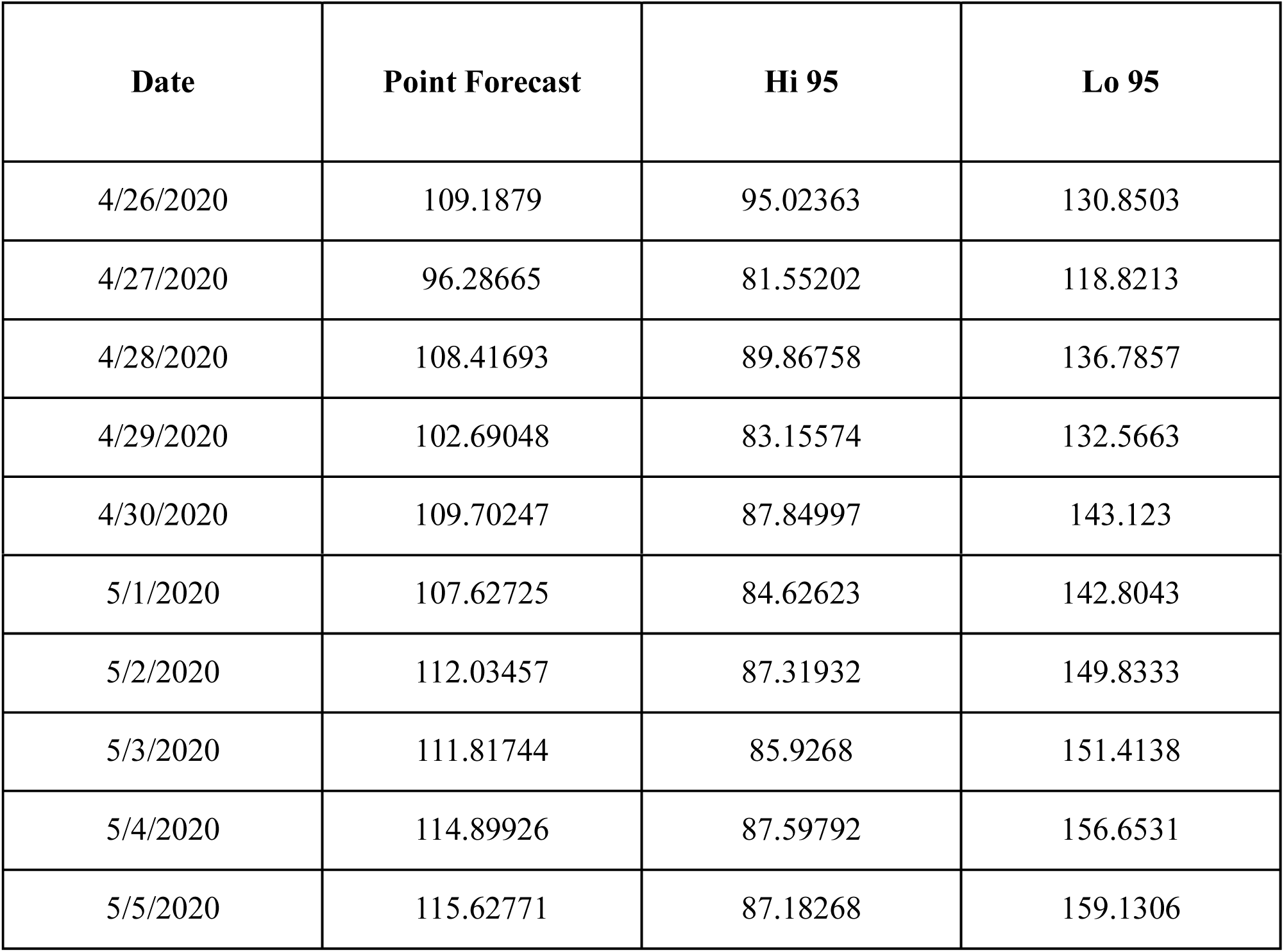
Future forecast values for COVID-19 cases in Nigeria

## Discussion

There was no case of COVID-19 in Nigeria until February of 2020. Since then, COVID-19 cases have been reported daily by the NCDC and on an uptrend. Following the current global strategies aimed at limiting the spread of the Coronavirus, most countries, including Nigeria, have restricted movements and asked people to maintain social distancing and globally about 3 billion persons are currently being confined in one way or another. Due to high illiteracy and poverty rate in most developing countries, there is need to understand the trend of the spread of COVID-19 and extrapolate the implications of the strategies employed by the government in curbing the spread of the disease.

Time-series analysis is a method to extrapolate predictions, in which a mathematical model is established according to the regularity and trend of the observed historical values with time (L. Liu, Luan, Yin, Zhu, & Lü, 2016; Q. Liu, Liu, Jiang, & Yang, 2011) AND has been widely used in predicting the spread of infectious diseases in recent years. For instance, some scholars have used this model to predict the spread of TB, mumps, measles, encephalitis B, and hand-foot-and-mouth disease (Allard, 1998; Jiabing, 2007; JIN et al., 2008; Musa, 2015; Wu et al., 2008; Yue et al., 2015). These diseases are similar to COVID-19 in time distribution (Organization, 2020; Peiris et al., 2003; Sahin, 2020). In this study, applying time series models as described above, to study the trend of the Covid-19 pandemic in Nigeria. Candidate models were obtained using the autocorrelation function (ACF) and the partial autocorrelation function (PACF), models were formulated based on the spikes observed from the ACF and PACF chart (Figure 2).

ARIMA (1,1,0) was confirmed to be the optimal model based on the lowest AICc value. This model was then applied to study the trend of COVID-19 from April 26, 2020 to May 5, 2020. The incidence of COVID-19 from April 26 to May 5, 2020, shows an increasing growth steep in Nigeria (115.62771, 95% confidence limit of 87.18268-159.1306), which peak cannot be said to have been seen or reached yet. According to these results, we must be alert of the possibilities of COVID-19 spreading more than its currently observed as seen from the models’ prediction. The proposed ease of lockdown restrictions by the federal government is expected to commence on May 4, 2020, and there is no indication of the trend to expect following the ease on restrictions. Countries like Germany have experienced an increased number in Covid19 positive cases after easing the country’s restrictions. Based on our study’s result, the trend of the spread of COVID-19 in Nigeria is expected to move in a upward trend. Having established a suitable model, Nigeria can apply this model to predict the trend of COVID-19 in the country.

## Conclusion

Government through the constituted presidential task force should be conscious of the wider spread if nothing is done as an addition to current lockdown for it is evident that the lockdown is not reducing the COVID-19 spread as expected.

## Data Availability

Data used from the Nigeria center for Disease Control (NCDC) is free and available for public use

